# Improving Individualized Rhabdomyosarcoma Prognosis Predictions Using Somatic Molecular Biomarkers

**DOI:** 10.1101/2024.09.04.24313032

**Authors:** Mark Zobeck, Javed Khan, Rajkumar Venkatramani, M. Fatih Okcu, Michael E. Scheurer, Philip J. Lupo

**Affiliations:** Baylor College of Medicine, Department of Pediatrics, Division of Hematology/Oncology, Houston, Texas; Texas Children’s Hospital, Texas Children’s Cancer and Hematology Centers, Houston, Texas; Center for Cancer Research, National Cancer Institute, Bethesda, Maryland

## Abstract

**Purpose:** Molecular markers, such as *FOXO1* fusion genes and *TP53* and *MYOD1* mutations, increasingly influence risk-stratified treatment selection for pediatric rhabdomyosarcoma (RMS). This study aims to integrate molecular and clinical data to produce individualized prognosis predictions that can further improve treatment selection.

**Patients and Methods:** Clinical variables and somatic mutation data for 20 genes from 641 RMS patients in the United Kingdom and the United States were used to develop three Cox proportional hazard models for predicting event-free survival (EFS). The ‘Baseline Clinical’ (BC) model included treatment location, age, fusion status, and risk group. The ‘Gene Enhanced 2’ (GE2) model added *TP53* and *MYOD1* mutations to the BC predictors. The ‘Gene Enhanced 6’ (GE6) model further included *NF1*, *MET*, *CDKN2A*, and *MYCN* mutations, selected through LASSO regression. Model performance was assessed using likelihood ratio (LR) tests and optimism-adjusted, bootstrapped validation and calibration metrics.

**Results:** The GE6 model demonstrated superior predictive performance, offering 39% more predictive information than the BC model (LR p<0.001) and 15% more than the GE2 model (LR p<0.001). The GE6 model achieved the highest discrimination with a C-index of 0.7087, a Nagalkerke R^2^ of 0.205, and appropriate calibration. Mutations in *TP53*, *MYOD1*, *CDKN2A*, *MET*, and *MYCN* were associated with higher hazards, while NF1 mutation correlated with lower hazard. Individual prognosis predictions varied between models in ways that may suggest different treatments for the same patient. For example, the 5-year EFS for a 10-year-old patient with high-risk, fusion-negative, *NF1*-positive disease was 50.0% (95% confidence interval: 39-64%) from BC but 76% (64-90%) from GE6.

**Conclusion:** Incorporating molecular markers into RMS prognosis models improves prognosis predictions. Individualized prognosis predictions may suggest alternative treatment regimens compared to traditional risk-classification schemas. Improved clinical variables and external validation are required prior to implementing these models into clinical practice.

## Introduction

Rhabdomyosarcoma (RMS) is an aggressive soft tissue sarcoma with highly variable survival outcomes. RMS is the most common soft tissue sarcoma in children in the United States (US). Approximately 350 children and 150 adults develop RMS each year.^1^ Successful treatment in most cases may require any combination of surgical resection of the primary tumor, radiation to the primary and metastatic sites, and multiagent chemotherapy that can last anywhere between 24 and 48 weeks. Prognostic clinicopathological features determine the intensity of the treatment regimen. These features stratify cases into three primary risk groups in the US with widely varying survival reported within each group. Low-risk (LR) RMS consistently demonstrates 5-year event-free survival (EFS) ≥90%, intermediate-risk (IR) RMS has a 5-year EFS between 50-85%, and high-risk (HR) RMS has a 5-year EFS between 5-45%.^2^

Molecular biomarkers have been shown to improve RMS prognostic stratification. *FOXO1* gene fusions with either *PAX3* [t(2;13)] or *PAX7* [t(1;13)] have significant prognostic implications and have been incorporated into standard risk-stratified treatment assignments in the United States.^3, 4^ More recently, tumors with mutations in *MYOD1* or *TP53* genes have been shown to have worse prognoses.^5^ These have been incorporated into the current Children’s Oncology Group (COG) LR clinical ARST2032 to increase the treatment intensity for patients with these mutations and to enrich a subset of very low-risk patients who are negative for the mutations to test the effect of treatment de-escalation.^6^ These molecular markers have improved the accuracy of prognosis classification, thereby enabling progress in studying treatments.

Despite these successes, improvements in RMS risk stratification are still needed. The prevalence of mutations in either *MYOD1* or *TP53* among *PAX::FOXO1* fusion-negative (FN) RMS has been reported to be around 3% and 13%, respectively.^5^ No other biomarkers have been clearly identified among FN RMS. Nor are there established biomarkers for *PAX::FOXO1* fusion-positive (FP) RMS. The most common alterations among FP RMS are amplifications of *MYCN* at 10% prevalence and *CDK4* at 13%. Studies suggest these lesions have negative prognostic significance, while other analyses have been equivocal.^5, 7, 8^ Given the wide range of survival outcomes between and within risk strata, new biomarkers can enhance risk-adapted treatment selection by identifying patients who may safely receive dose-reduced therapy or may benefit from treatment intensification.

The aim of this exploratory study is to identify new predictive biomarkers that can improve RMS risk stratification. To accomplish this aim, we leveraged a large dataset of over 600 pediatric patients for whom a panel of somatic mutation data was available to develop statistical models that combine clinical and genetic variables to predict 5-year event-free survival (EFS). By focusing on prediction, we quantified the amount of prognostic information for promising biomarkers and evaluated how prognoses change according to mutation status.

## Methods

### Study Population & Data Collection

We conducted a retrospective analysis of pediatric rhabdomyosarcoma (RMS) cases from the United Kingdom (UK) and the United States (US). Case data were obtained from a publicly available dataset hosted by the National Institutes of Health OncoGenomics data portal.^5, 9^ Case-level clinical data for age at diagnosis, risk group, fusion status, and treatment location (UK or US) were available. Several variables were modified to harmonize the datasets (see the supplement of Shern et al., 2021 for full details^5^). A dedicated *FOXO1* fusion assay was unavailable for 40 of the 126 cases designated as alveolar histology after central pathology review. These cases were presumed to be positive for *PAX::FOXO1* fusions. The risk group designation was modified to accommodate differences in treatment protocols between the treatment contexts in the UK and US. Low risk was defined as non-metastatic embryonal or FN RMS with an orbital, paratesticular, female genitourinary, or head/neck (non-parameningeal) primary site. High risk was defined by the presence of any metastatic disease. Intermediate risk was defined as not meeting the definition of high or low risk.

### Somatic Mutation Data

Somatic mutation data for 39 genes were generated for tumor samples using next-generation sequencing as described in the supplement of Shern et al., 2021.^5^ Only mutations with a frequency of five or more in the cohort were retained for final analysis, resulting in 20 total genes evaluated. Nineteen genes had four or fewer mutations.

### Model Development

The primary outcome to predict was 5-year EFS where relapse, disease progression, second malignant neoplasm, and death were classified as events. We developed three Cox proportional hazard models:

1. **Baseline Clinical (BC) Model:** Incorporated age, *PAX::FOXO1* fusion status, risk group, and treatment location as predictors.
2. **Gene Enhanced 2 (GE2) Model:** Included all predictors from BC and added variables for *TP53* and *MYOD1* mutations.
3. **Gene Enhanced 6 (GE6) Model:** Included all predictors from GE2 and added variables for *CDKN2A*, *MET, MYCN*, and *NF1* mutations.

### Predictor Selection & Model Specification

The variables in BC were chosen to reflect the standard variables used in prognostic stratification for treatment assignment. Tumor stage and clinical group were unavailable in the complete dataset. The modified risk grouping designed to harmonize the datasets from the US and UK also functioned as a proxy variable to reflect standard prognostic categories. Due to considerations that there may be a nonlinear relationship between age and outcome, the coefficient for age was evaluated using a restricted cubic spline with four knots. All genetic variants were dichotomous variables specifying whether the variant was present or absent. *TP53* and *MYOD1* were included in the GE2 model to evaluate the added predictive value of these mutations that have been newly implemented into risk-stratification schemas in the ongoing COG trial ARST2032.

Predictors for the GE6 model were selected using least absolute shrinkage and selection operator (LASSO) Cox regression, a form of regularized regression that applies a penalty term to the coefficients to enable the selection of the most informative predictors while avoiding overfitting to the training data. The four variables from BC were forced to be included in the GE6 model. The candidate variables included the 20 candidate genes, a variable for the total number of mutations in an individual tumor, and a variable for the number of RAS pathway mutations (mutations in *NRAS*, *KRAS*, *HRAS*, *PIK3CA*, *NF1*, or *FGFR4*). The penalty term for the LASSO model was selected through cross-validation to identify the value that maximized the concordance index with the least number of variables. An unregularized Cox regression model was then trained using the variables selected by this procedure to produce the final version of the GE6 model. Due to the possibility of unstable variable selection by LASSO and the risk of overfitting when using unregularized coefficients, we evaluated the entire model development process using the bootstrap procedure and demonstrated that it preserved the desired performance characteristics to propose new predictive biomarkers (Appendix A, Section 1).

### Internal Validation, Model Comparison, & Analysis

Model performance for all three models was assessed and compared across a range of metrics. The likelihood ratio test and the percent of added information (one minus the ratio of the variance of outcome predictions for the smaller model over the variance of predictions from the larger model) were used for comparisons of the overall improvements in predictive performance with the addition of the molecular variables to the models. C-indices, Nagalkerke’s R^2^, calibration slopes, and Gini’s mean difference measures were calculated and corrected for optimism (a form of internal validation) using the bootstrap procedure. Apparent and bias-corrected calibration curves for 5-year EFS were also produced through bootstrap resampling. The final model performance was described by reporting the hazard ratios for the predictors, individual survival predictions, and the change in predicted survival probabilities for the same observation between two models. All statistical analyses were performed using R version 4.3.1 using the “tidyverse,” “gtsummary,” “survival,” “survminer,” and “rms” packages.^10–15^

## Results

Of the 641 patients in the dataset, nine were excluded due to missing values in age, fusion status, or risk group and 632 were eligible for analysis. Table 1 lists the demographic and clinical variables for the entire dataset and by treatment cohort (COG or UK). Two hundred and seventeen (36%) patients in the dataset experienced an event. The median time to event was 445 days (Interquartile range 18 – 1190 days). The median follow-up time among censored patients was 2,691 days (565 – 5,461 days).

**Table 1.** Demographic characteristic of the patient cohort overall and by treatment context.

| Characteristic | All Patients<br>N = 632<sup>1</sup> | Treatment context |  |
| --- | --- | --- | --- |
|  |  | COG<sup>1</sup>, N = 344<sup>1</sup> | UK<sup>2</sup>, N = 288<sup>1</sup> |
| Sex, n (%) |  |  |  |
| Male | 416 (66%) | 232 (67%) | 184 (64%) |
| Female | 216 (34%) | 112 (33%) | 104 (36%) |
| Age, med (Q1, Q3)<sup>3</sup> | 6.0 (3.1, 11.4) | 6.4 (3.4, 12.2) | 5.3 (2.8, 9.5) |
| Risk group, n (%) |  |  |  |
| Low | 220 (35%) | 124 (36%) | 96 (33%) |
| Intermediate | 298 (47%) | 147 (43%) | 151 (52%) |
| High | 114 (18%) | 73 (21%) | 41 (14%) |
| FOXO1 Fusion, n (%) |  |  |  |
| Negative | 508 (80%) | 275 (80%) | 233 (81%) |
| Positive | 124 (20%) | 69 (20%) | 55 (19%) |
| Total mutations, n (%) |  |  |  |
| 0 | 184 (29%) | 88 (26%) | 96 (33%) |
| 1 | 247 (39%) | 136 (40%) | 111 (39%) |
| 2 | 144 (23%) | 85 (25%) | 59 (20%) |
| 3 | 57 (9.0%) | 35 (10%) | 22 (7.6%) |
| *TP53* mutation, n (%) | 74 (12%) | 37 (11%) | 37 (13%) |
| *MYOD1* mutation, n (%) | 17 (2.7%) | 11 (3.2%) | 6 (2.1%) |
| *CDKN2A* mutation, n (%) | 23 (3.6%) | 17 (4.9%) | 6 (2.1%) |
| *NF1* mutation, n (%) | 79 (13%) | 41 (12%) | 38 (13%) |
| *MYCN* mutation, n (%) | 13 (2.1%) | 10 (2.9%) | 3 (1.0%) |
| *MET* mutation, n (%) | 9 (1.4%) | 6 (1.7%) | 3 (1.0%) |
| Event<sup>4</sup> | 236 (37%) | 121 (35%) | 115 (40%) |
| Death | 187 (30%) | 90 (26%) | 97 (34%) |

<sup>1</sup>COG, Children's Oncology Group. <sup>2</sup>UK, United Kingdom. <sup>3</sup>Med, median; Q1, quartile 1; Q3, quartile 3.
<sup>4</sup> Events were defined as disease progression or relapse, second malignant neoplasm, or death.

The GE6 model demonstrated the best overall predictive performance ability. By the likelihood ratio test, the GE6 model demonstrated superior predictive performance compared to the BC (χ^2^ = 71.4 on 6 degrees of freedom [df], *p* < 0.001) and GE2 (χ^2^ = 29.3 on 4 df, *p* < 0.001) models. The GE2 model similarly demonstrated superior predictive ability compared to BC (χ^2^ = 42.2 on 2 df, *p* < 0.001). The GE6 model provided 39% more predictive information than the BC model and 15% more than the GE2 model. GE2 provided 28% more information than the BC model.

Across all optimism-corrected performance metrics, the GE6 model performed better than GE2, which performed better than BC (Table 2). GE6 achieved the highest discrimination of all the models with a C-index of 0.7087. All three models demonstrated good bias-corrected calibration curves for 5-year EFS, with only slight tendencies toward regression to the mean for each model upon bias correction (slope of the blue line < 1, Figure 1; regression slope metric < 1, Table 2).

**Figure 1.**
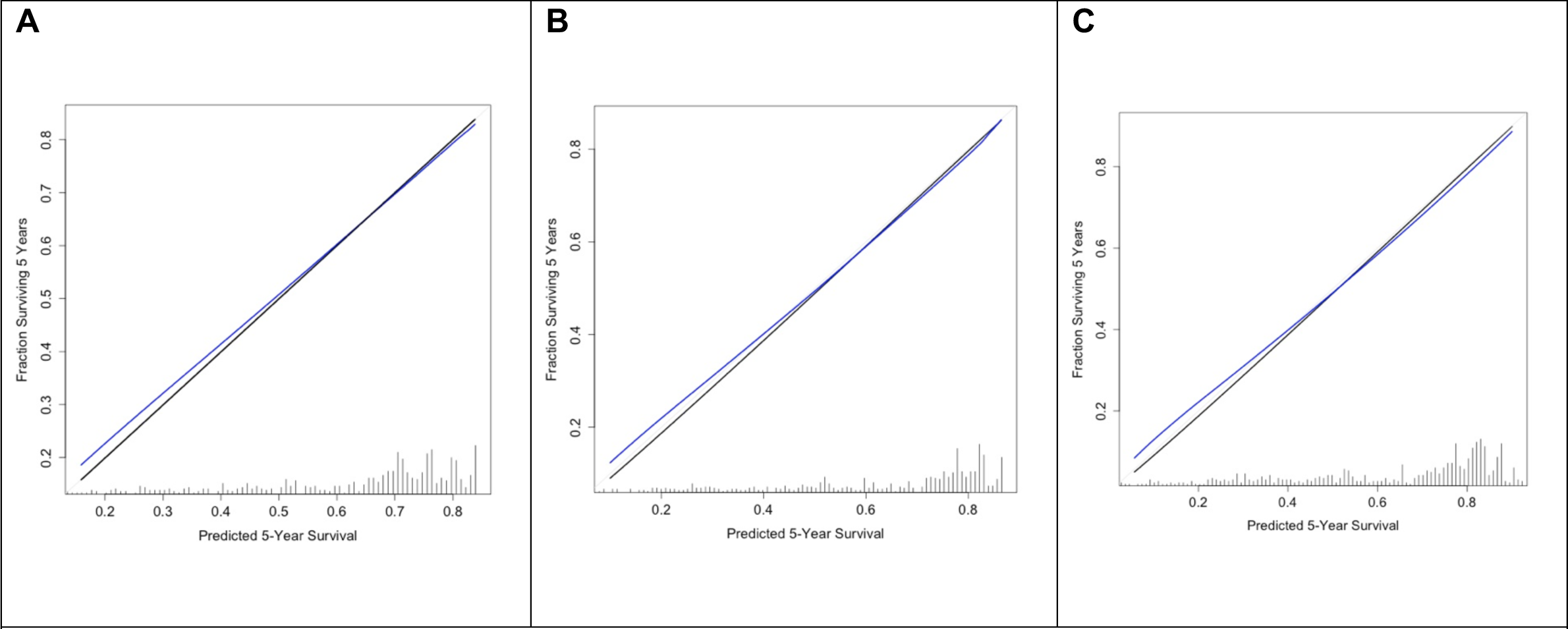
Apparent (black line) and bias-adjusted (blue line) calibration curves for. A) the Baseline Clinical model, B) the Gene Enhanced 2 model, and C) the Gene Enhanced 6 model. The gray diagonal line represents perfect calibration. Tick marks at the bottom represent the relative distribution of predicted probabilities in the dataset.

**Table 2.** Bootstrapped optimism-corrected performance metrics for each prediction model.

| **Metric** | **BC** | **GE2** | **GE6** |
| --- | --- | --- | --- |
| Concordance index | 0.6512 | 0.6905 | 0.7087 |
| Nagalkerke's $R^2$ | 0.1236 | 0.1780 | 0.2046 |
| Calibration slope | 0.9369 | 0.9403 | 0.9087 |
| Gini's mean difference | 0.6379 | 0.7864 | 0.8704 |

BC, Baseline Clinical Model; GE2, Gene Enhanced 2 model; GE6, Gene Enhanced 6 model.

For the GE6 model, all variables, including the nonlinear component for age, were significantly associated with EFS by the Wald test (Table S3-4 for log-hazard coefficient values). The hazard ratios for the categorical variables are presented in Table 3. Mutations were associated with a higher hazard for all genes except for NF1, which was associated with a lower hazard. The hazard for age showed a U-shaped relationship (Figure 2), with younger and older ages showing higher log relative hazard compared to around 5 years. The variables in BC and GE2 showed similar patterns to GE6. Variable Wald tests, coefficient values, logarithmic hazard, and hazard ratios are provided in Tables S3-6.

**Figure 2.**
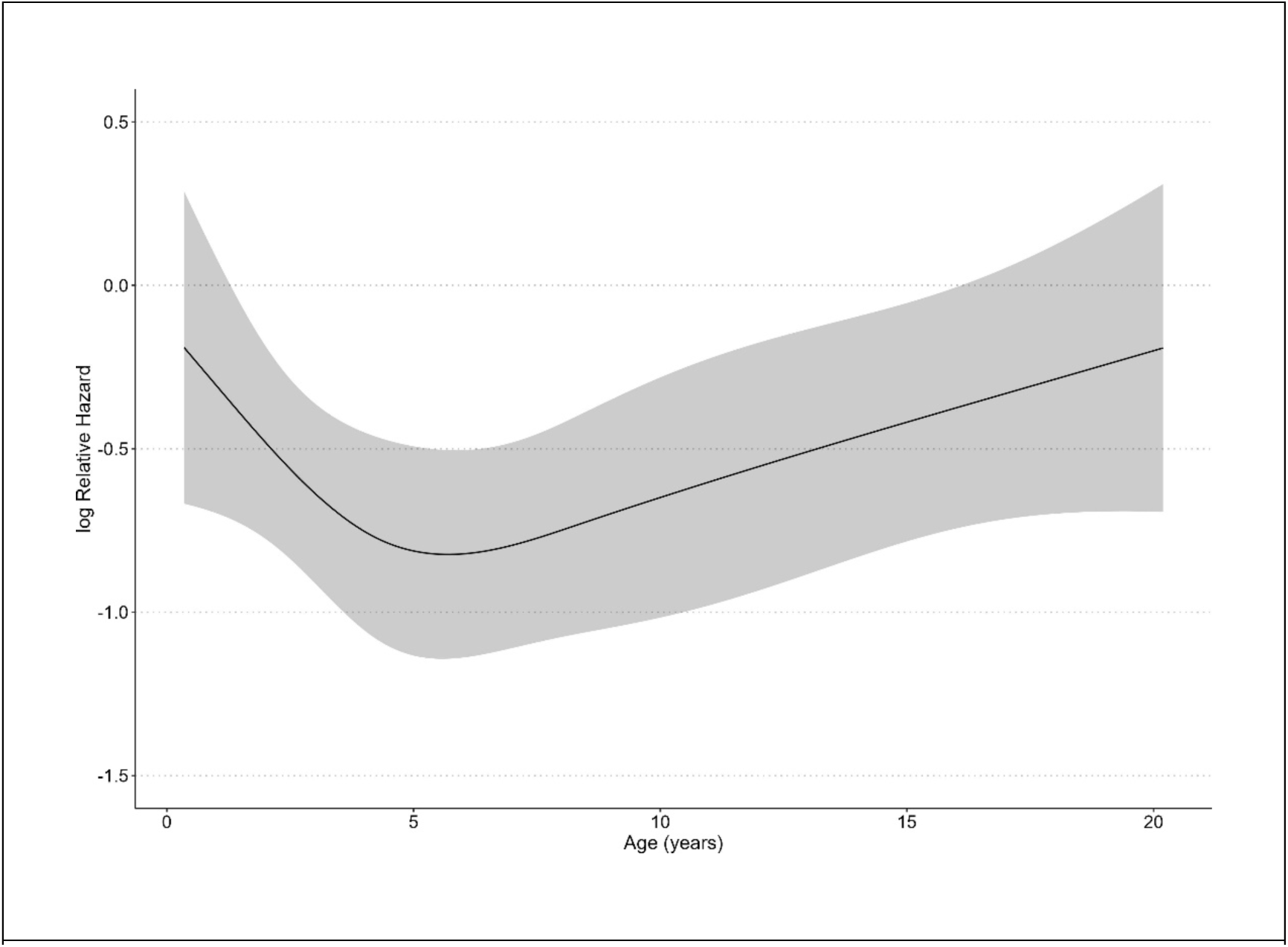
Estimates of log relative hazard values by age from the Gene Enhanced 6 model. The curve shows a flexible fit due to the use of a restricted cubic spline to capture nonlinear relationships between age and 5-year EFS. The black line represents the log relative hazard point estimate by age. The gray ribbon represents the 95% confidence interval.

**Table 3.** Hazard ratios for the categorical variables in the Gene Enhanced 6 model.

| Variable | Estimate | 95% Confidence Interval | *P* value |
| --- | --- | --- | --- |
| Risk Group (reference: Low) |  |  |  |
| Intermediate | 1.31 | 0.91 - 1.91 | 0.130 |
| High | 3.20 | 2.12 - 4.82 | < 0.001 |
| *PAX::FOXO1* fusion (+) | 2.43 | 1.72 - 3.43 | < 0.001 |
| Treated in UK | 1.49 | 1.14 - 1.95 | 0.007 |
| *MYOD1* mutation | 4.61 | 2.44 - 8.70 | < 0.001 |
| *TP53* mutation | 2.66 | 1.83 - 3.85 | < 0.001 |
| *CDKN2A* mutation | 3.22 | 1.74 - 5.93 | < 0.001 |
| *MET* mutation | 5.98 | 2.55 - 13.98 | < 0.001 |
| *MYCN* mutation | 2.01 | 1.02 - 3.95 | 0.048 |
| *NF1* mutation | 0.56 | 0.34 - 0.92 | 0.020 |

Survival predictions for each patient in the cohort varied significantly across models. Figure 3 demonstrates how survival predictions changed for the patients from BC compared to GE2 (left panel) and GE6 (right panel). The figure shows that including genetic information results in very different survival predictions for some patients. Table 4 provides survival predictions for specific types of patients according to the BC and GE6 models. Predictions from BC, where genetic information is ignored, may be compared to predictions from GE6 for patients with and without the specified mutations. Table 4 shows that a 7-year-old patient with low-risk, FN disease who is positive for a *TP53, MYOD1, CKDN2A,* or a *MET* mutation have lower expected survival when mutation information is included (right column) compared to patients without those mutations (middle column) or to when genetic information is ignored (left column). As *MYCN* mutations were only observed in FP patients, survival predictions were produced for a 7-year-old patient with intermediate-risk, FP disease. Predictions for a 10-year-old with high-risk, FN disease were used for *NF1* mutations to demonstrate how prognosis improves given the mutation. For all predictions, 95% confidence intervals (CIs) are provided to demonstrate the range of survival predictions compatible with the data and model.

**Figure 3.**
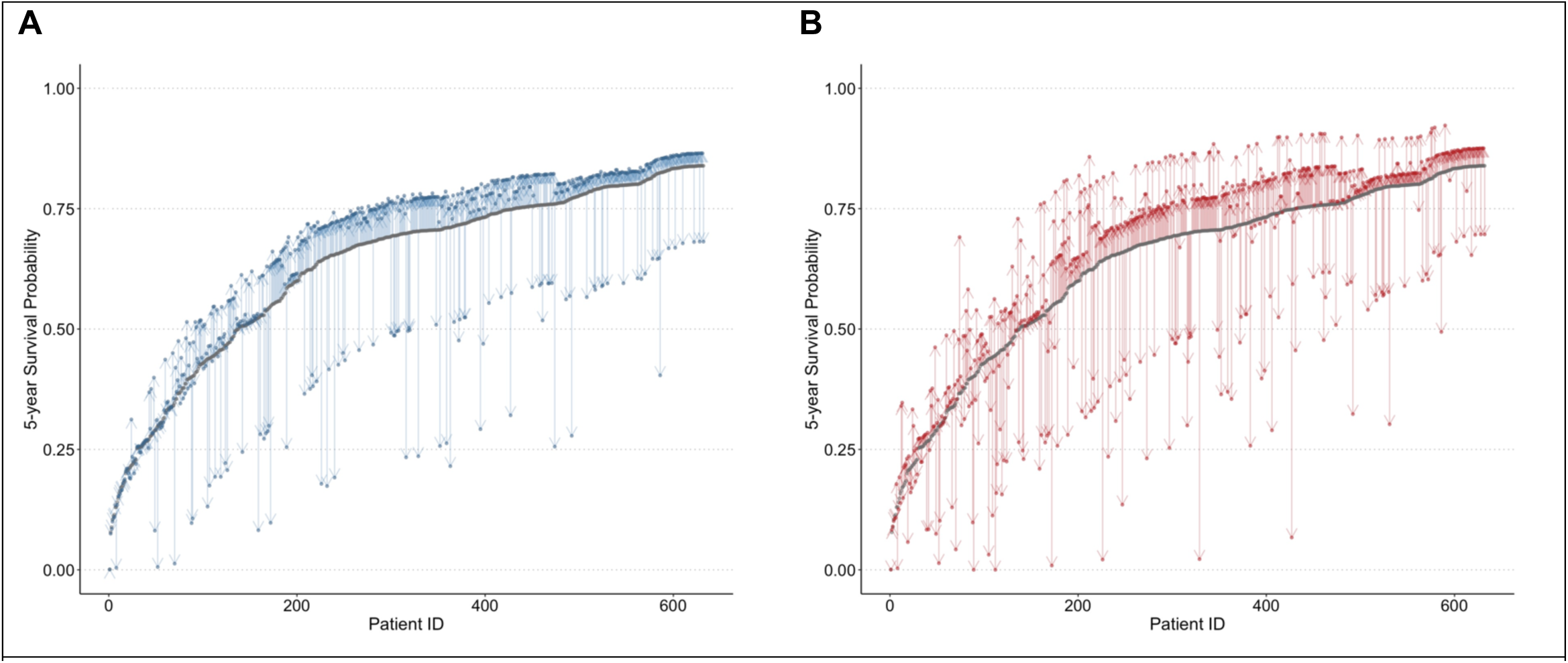
Predictive probability of surviving five years without an event for each patient in the dataset for. A) the GE2 model (blue) compared to BC (gray) and B) the GE6 model (red) compared to BC (gray). Points represent the point estimate for 5-year survival. Patients are aligned by lowest to highest estimated 5-year survival according to the BC model. Vertical arrows at the same Patient ID represent the difference in the estimated 5-year survival between the models. Both the GE2 and GE6 models estimate worse and improved survival chances relative to the BC model (panels A and B, colored compared to gray points).

**Table 4.** Predicted 5-year event-free survival from the BC and GE6 models for different types of patients.

| Patient Characteristics | BC | GE6 |  |
| --- | --- | --- | --- |
|  | Mutation Ignored | Mutation Negative | Mutation Positive* |
|  | 5-yr EFS<br>Mean (95% CI) | 5-yr EFS<br>Mean (95% CI) | 5-yr EFS<br>Mean (95% CI) |
| 7 yr, LR, FN |  |  |  |
| ***TP53*** | 84% (78-89%) | 87% (83-92%) | 69% (58-82%) |
| ***MYOD1*** | 84% (78-89%) | 87% (83-92%) | 53% (34-84%) |
| ***CDKN2A*** | 84% (78-89%) | 87% (83-92%) | 65% (49-86%) |
| ***MET*** | 84% (78-89%) | 87% (83-92%) | 36% (22-90%) |
| 7 yr, IR, FP |  |  |  |
| ***MYCN*** | 59% (49-71%) | 64% (54-76%) | 42% (23-75%) |
| 10 yr, HR, FN |  |  |  |
| ***NF1*** | 50% (39-64%) | 60% (49-74%) | 76% (64-90%) |

BC, Baseline Clinical Model; GE6, Gene Enhanced 6 model; CI, confidence interval; yr., year; LR, low risk; IR, intermediate risk; HR, high risk; FN, *FOXO1* fusion negative; FP, *FOXO1* fusion positive.
\*Mutations for all other genetic variables in GE6 are negative except for the gene indicated corresponding row.

## Discussion

### Overall Findings

Our results demonstrate that known and new molecular markers can improve prognosis predictions for RMS. We showed how mutation information *TP53* and *MYOD1,* known to be associated with survival, can improve prognosis predictions over predictions using only age, risk category, fusion status, and treatment location. We also described the added predictive utility of a new set of genes, *CDKN2A, MET, MYCN,* and *NF1.* We showed that each of these genes is significantly associated with 5-year EFS on multivariable modeling and that this set of genes improves the discrimination of predictions while maintaining adequate calibration. If the predictive potential of these new molecular markers is validated in an external cohort, they can be used to make risk stratification more accurate, as with *TP53* and *MYOD1,* and improve risk-adapted therapy assignment.

By predicting 5-year EFS for individual patients in Table 4, we demonstrated that expected survival can change dramatically in ways that may suggest alternative therapies when molecular markers are included. For low-risk disease, mutations in *TP53, MYOD1, CKDN2A,* and *MET* were associated with a decreased mean 5-year EFS that more closely resembled the prognosis of intermediate-risk patients of around 50-70%. *MYCN* amplification, observed exclusively in FP patients, was associated with a lower mean survival of less than 50%, similar to that of high-risk patients. By contrast, an *NF1* mutation in a high-risk patient was associated with an improved survival of 76%. In each scenario, the mean predicted survival changed to suggest an alternative risk category for the patient. This change in risk category, in turn, suggests that a change in treatment intensity may also be appropriate.

### New Molecular Markers

*CKDN2A, MET, MYCN,* and *NF1* all participate in a common genetic pathway characterized by the activity of several receptor tyrosine kinases (RTKs), and *RAS* and *PIK3CA* genes.^16^ *PAX::FOXO1* translocations represent important driver mutations in this pathway that upregulate the expression of MYCN, which increases the malignant transformation of RMS precursor cells.^17^ *CKDN2A, MET,* and *NF1* are commonly observed as driver mutations among FN disease.^16^ Although the mutation panel of the primary study emphasized genes within the RTK/*RAS*/*PIK3CA* axis, the identification of independently prognostic mutations lends support to the hypothesis that derangements along this axis lead to different clinical phenotypes of RMS beyond the primary FN/FP divide.

*NF1* (neurofibromin 1) plays an inhibitory role among genes in the *RAS* pathway and has been implicated in multiple cancers and cancer predisposition syndromes.^18^ *NF1* was the most commonly mutated gene among the candidate predictors, with 79 cases. It was associated with improved survival in this study (hazard ratio: 0.55 [0.34-0.91]) and resulted in a large improvement in predicted 5-year EFS from 50% (BC model) to 76% (GE6) for a 10-year-old patient with HR, FN RMS. Among *TP53* mutation*-*negative but *NF1* mutation-positive cases, the observed 5-year OS was 96% for the 22 low-risk cases (compared to 88% for the 173 *TP53* negative, *NF1* negative cases) and 86% for the 37 intermediate-risk cases (compared to 70% for 224 *TP53* negative, *NF1* negative cases). Similar survival outcomes were recently reported in a cohort of 14 *TP53* negative*, NF1* positive cases, which had a 5-year OS of 90%.^19^ Taken together, this is compelling evidence that *NF1* mutation could be a positive prognostic marker that warrants further evaluation.

*MET* (mesenchymal epithelial transition) is an oncogene that codes for a receptor tyrosine kinase (RTK) that impacts cellular survival, migration, and invasion that has been implemented as a pathogenic driver in a variety of cancer types.^6,8^ Expression of the RTK that it encodes, c-MET, has been previously associated with OS in a small case series of patients with ERMS,^20^ and downregulation of the MET receptor has been shown *in vitro* to result in decreased migration and metastatic behavior of RMS cells.^21^ Importantly, multiple therapies are available that target the c-MET protein.^22^ *MET* mutations yielded a hazard ratio of 5.91 (2.53-13.83) and resulted in a large decrease in predicted survival from 84% (BC model) to 36% (GE6) for a 7-year-old LR, FN patient, although the confidence interval for this estimate was large due to the small number of cases in the dataset. Nonetheless, these results suggest that *MET* mutations may be a rare but important targetable prognostic marker that deserves further evaluation.

*CDKN2A* (cyclin-dependent kinase inhibitor 2A) is a tumor suppressor gene that regulates cell cycle progression.^18^ It has been previously associated with poor prognosis for soft tissue sarcomas.^23^ The adjusted hazard ratio from the GE6 model for a mutation was 3.18 (1.72-5.87), and the decrease in the predicted survival was comparable to *MYOD1* and *TP53* mutations, although there was a large amount of residual uncertainty in the confidence intervals. These results support the continued evaluation of *CKDN2A* as a prognostic biomarker.

*MYCN* codes for a transcription factor that regulates a variety of cellular processes. Amplification of this gene has been implicated in the development of neuroblastoma and other cancers.^18, 24^ *MYCN* is most commonly amplified in FP disease and was observed exclusively in FP disease in this cohort.^5, 16^ Our results indicated that *MYCN* is associated with 5-year EFS independent among FP cases independently from the other variables in GE6 with an adjusted hazard ratio of 1.97 (1.00-3.88). However, this finding should be considered more tenuous than the other identified genes, as the association was comparatively smaller, the *p-*value was larger, and the selection of this gene in the final model on bootstrap evaluation of the LASSO procedure was the most inconsistent (selected in 65% of bootstrap samples).

### Established Molecular Markers

This study provides supportive evidence to reinforce the importance of *TP53* and *MYOD1* mutations as molecular markers of poor prognosis. Survival rates for *MYOD1* reported in small case series range between 0-30%, even for low-risk disease.^25–27^ In the primary analysis of this cohort, Shern et al. demonstrated that the *MYOD1* mutation had an overall five-year EFS of 18%, and had a hazard ratio after adjusting for risk group of 5.58 (95%CI: 2.80-11.2),^5^ which decreased to 4.57 (2.41-8.62) after adjustment with GE6. Our analysis complements the original analysis by quantifying how survival predictions change when *MYOD1* mutation status is considered. Of note, the mean prognosis predictions for patients classified as low-risk are higher than the survival for low-risk diseases reported in the literature. However, the 95% CI for the predictions is wide and includes values in the low 30%. Direct comparison between results from other studies and the present study is difficult because the predictions are adjusted for age and treatment cohort, and the risk groupings in this study are non-standard in order to harmonize the US and UK data. If there is miscalibration, it may be due to the small number of cases preventing precise risk estimation after adjusting for age, risk group, and other covariates. Alternatively, *MYOD1* mutation may confer a negative prognosis across risk groups. Therefore, interactions between risk groups and mutation status should be assessed in future modeling efforts. Nevertheless, this study supports the clear associations between *MYOD1* and poor prognosis.

In the primary analysis of *TP53* mutations in this cohort, Shern et al. reported a hazard ratio after adjusting for risk group for *TP53* of 1.97 (1.13-3.44) and an overall 5-year EFS of 49%, which varied by risk group when stratified by treatment cohort.^5^ After adjustment in the GE6 model, the hazard ratio rose to 2.72 (1.88-3.93). The calibration of predictions from the models compares favorably to the observed risk-stratified survival rates in the primary analysis. Like *MYOD1*, these results quantify how survival predictions change due to including information about *TP53* mutation in predictions, reinforcing the evidence of its importance as a prognostic marker.

### Limitations & Future Directions

While the prediction models we developed performed well, the limitations of this study entail that further development is required prior to clinical implementation. The risk grouping variable used in the model represents a consensus grading to facilitate comparison between UK and US treatment contexts. To use these models in specific clinical contexts, the risk grouping variables must reflect the actual clinicopathological criteria used for prognostic stratification, such as the current staging system used in COG clinical trials. These limitations also impaired comparing the models to the standard risk-adapted treatment assignment schema. Treatments are also not included in predictions, which are important variables when predicting prognosis among patients who receive treatment.^28, 29^ Using the LASSO procedure to select the molecular predictors for GE6 can induce overfitting, making the predictors appear more predictive than they will be in a general population. While the bias-corrected calibration slope did not demonstrate major overfitting, the prognostic value of these predictors must be verified in a pre-specified model in an external cohort. If the performance of models with these improvements can be validated, then they will be fit for use to assist with prognosis-informed clinical decision-making.

### Conclusion

In a large cohort of pediatric patients with RMS, we identified several promising prognostic biomarkers that may improve risk-adapted treatment assignment. Mutations in *NF1* are associated with a substantially decreased risk of treatment failure, and mutations in *CDNK2A, MET, and MYCN* are associated with an increased risk. We also quantified the risk of treatment failure in the known prognostic markers *MYOD1* and *TP53*. Finally, we demonstrated that clinical prediction models that combine clinicopathological and molecular prognostic markers can adequately predict survival. With further development and external validation, these models could be implemented into clinical practice to improve how providers care for pediatric patients with RMS.

## Data Availability

The source data are available on the National Cancer Institute's Center for Cancer Research Oncogenomics data platform at https://clinomics.ccr.cancer.gov/clinomics/public.

## Acknowledgments

The authors wish to thank the team of Shern at al., who published the original dataset^5^ and the National Institutes of Health Oncogenomics team for hosting the data.

## Author Contributions

### Conception and design

Mark Zobeck, Philip J. Lupo, M. Fatih Okcu, Rajkumar Venkatramani, Michael E. Scheurer

### Financial support

Mark Zobeck

### Provision of study materials or patients

Javed Khan, Mark Zobeck

### Collection and assembly of data

Mark Zobeck, Javed Khan

### Data analysis and Interpretation

Mark Zobeck, Philip J. Lupo, M. Fatih Okcu, Rajkumar Venkatramani, Michael E. Scheurer, Javed Khan

### Manuscript Writing

Mark Zobeck, Philip J. Lupo, M. Fatih Okcu, Rajkumar Venkatramani, Michael E. Scheurer, Javed Khan

### Final Approval of Manuscript

Mark Zobeck, Philip J. Lupo, M. Fatih Okcu, Rajkumar Venkatramani, Michael E. Scheurer, Javed Khan

