## Supplementary Materials for "Improving Individualized Rhabdomyosarcoma Prognosis Predictions Using Somatic Molecular Biomarkers"

### 1 Section 1: Model Development

#### 1.1 Selection of the $\lambda$ parameter for LASSO regression

LASSO regression induces sparsity in the model by penalizing the sum of the absolute values of the coefficients. The penalty term is controlled by the $\lambda$ parameter. We used the `cv.glmnet` function to select the optimal $\lambda$ parameter for the LASSO regression model. The optimal $\lambda$ parameter was selected using 10-fold cross-validation using the Concordance index (C-index) as the optimization metric. The optimal $\lambda$ parameter was selected as the value that that maximized the C-index with the least number of variables (Figure S1). A penalty term for $log(\lambda) = -3.5$ was selected.

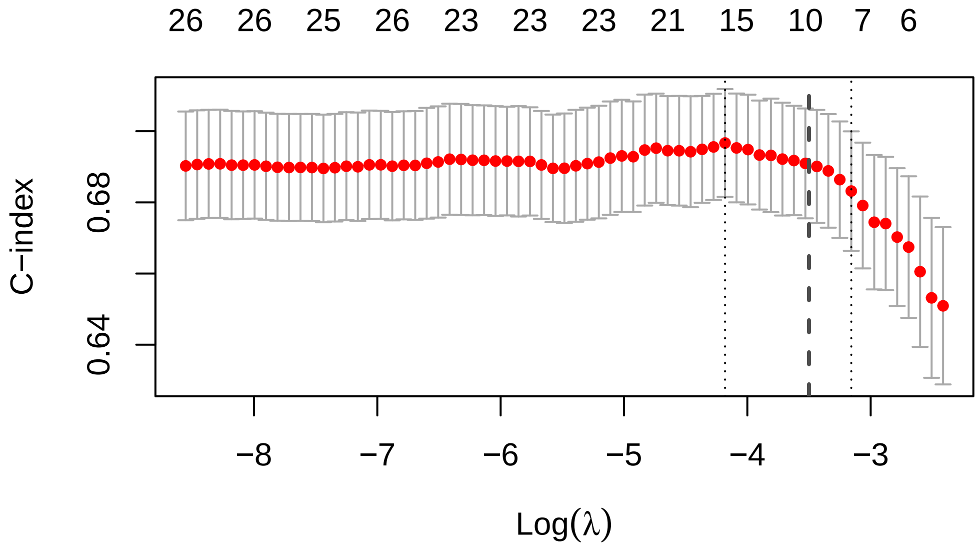

Figure 1: Cross-validated concordance index for LASSO regression model by log(lambda) values. The chosen log(lambda) value is indicated by the thick vertical dashed line. The left thin vertical dashed line represents the log(lambda) value that maximized the cross-validation C-indxex. The right dashed line represents the largest value of log(lambda) such that the mean cross-validated C-index is within one standard error of the minimum mean cross-validated C-index.

#### 1.2 Evaluating of the stability of variable selection for GE6

The selection of specific variables by LASSO regression is unstable. Small changes in the data can result in different sets of variables. To evaluate the stably the $log(\lambda) = -3.5$ penalty term selects variables, we used a bootstrap algorithm to simulate perturbations of the dataset. A LASSO Cox model using the penalty term was fit on a dataset the same size as the original dataset that was derived from sampling the original with replacement. The variables that were identified by this process were recorded. This procedure was then repeated 300 times, and the frequency of the selected variables was evaluated. This procedure demonstrated that the variables included in the GE6 model were each retained in greater than 200 out of 300 bootstrapped models. (Figure S1). *MYCN* was just above the threshold at 202 times (67.3%

of bootstrapped models). *CDKN2A*, *MET*, and *NF1* were selected in 87-89% of bootstraps. *MYOD1* and *TP53* were selected in greater than 99% of bootstraps.

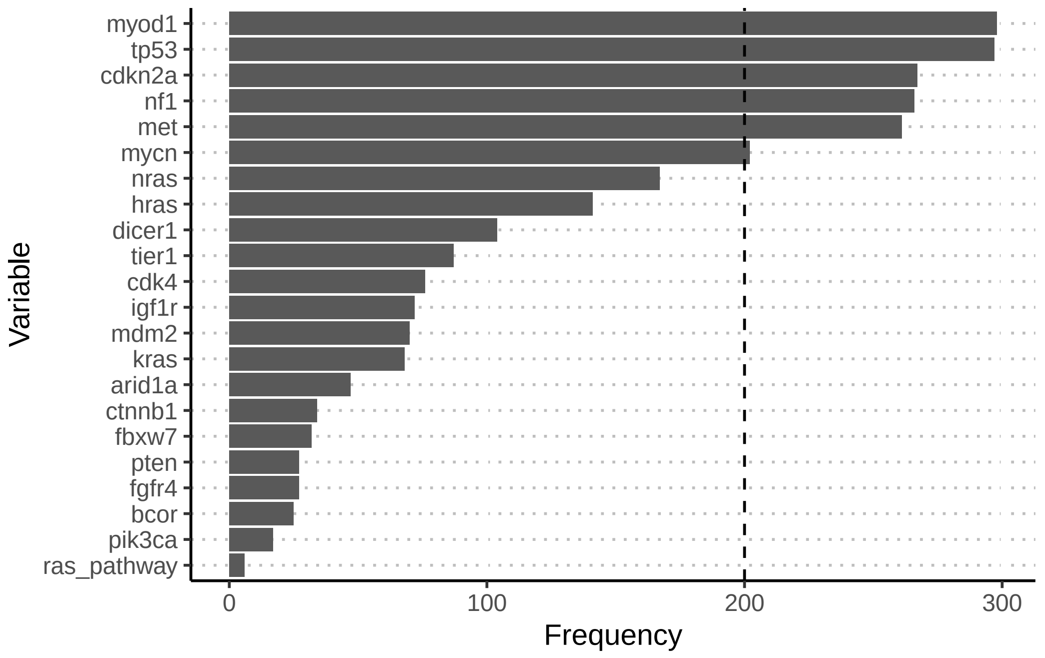

Figure 2: Frequency of variables selected in LASSO Cox Model. The dashed line represents the threshold of 200 selections in 300 bootstrap runs.

#### 1.3 Evaluating overfitting in unregularized coefficients

We chose to use an unregularized model to facilitate modeling age flexibly with restricted cubic splines and to produce confidence intervals on predicted survival. However, reporting unregularized coefficient values after the LASSO procedure may cause overfitting to the training data.

We evaluated the possibility of overfitting in the model development process in two ways.

The first evaluation was to perform bootstrap validation on the entire model fitting process and calculate optimism-adjusted C-index. Instead of using the unregularized Cox model to make predictions as in the main text, the LASSO model with regularized coefficients was used to predict outcomes and calculate the C-index. The steps in the procedure were:

1. **Extract Bootstrap Sample and Fit Model:**
    - Randomly sample the data with replacement to create a bootstrap sample.

- Fit a LASSO Cox model with $log(\lambda) = -3.5$ to the bootstrap sample using a predefined.

2. **Calculate C-Indexes:**
   - Predict risk for the bootstrap sample and calculate the c-index.
   - Predict risk for the original data and calculate the c-index.

3. **Run Bootstrapping Procedure:**
   - Perform 300 bootstrap iterations on the data, recording c-indexes for each iteration.

4. **Calculate Optimism:**
   - Estimate optimism by calculating the mean difference between the c-indexes from the bootstrap samples and the original data.

5. **Fit Final Model and Optimism-Corrected C-Index:**
   - Fit the LASSO Cox model to the entire dataset and calculate the apparent C-index.
   - Subtract the optimism from the apparent C-index to yield the optimism-corrected C-index.

The final result was an optimism-corrected C-index of for the LASSO Cox model of 0.7011. The optimism was 0.00384. This value was close to the optimism-corrected C-index of the unregularized version of the model of 0.7087 and the optimism was 0.0124. As expected, the optimism was slightly higher in the unregularized version, although it was also small. More importantly, the bias corrected performance was slightly higher, likely owing to the flexibility of modeling using restricted cubic. Overall, these results demonstrate that the LASSO Cox model and the unregularized Cox model had similar performance in this dataset. Given the advantages of the unregularized version, it was chosen as the final model form.

The second evaluation of overfitting was calculating the optimism-correct calibration slope and producing flexible, bias-corrected calibration curves for the GE6 model (Table 2 and Figure 1 of the main text). Each of these demonstrated that using the unregularized coefficients in this dataset induced minimal additional overfitting and miscalibration when corrected for optimism.

### 2 Section 2: Additional Modeling Results

#### 2.1 Gene Enhanced 6 Model

##### 2.1.1 Coefficients

| Variable | Chi-Square | d.f. | P |
|---|---|---|---|
| age | 8.27 | 3 | 0.04 |
| Nonlinear | 7.21 | 2 | 0.03 |
| risk_group | 40.17 | 2 | 0.00 |
| fusion | 25.29 | 1 | 0.00 |
| cohort | 7.33 | 1 | 0.01 |
| myod1 | 21.92 | 1 | 0.00 |
| tp53 | 28.07 | 1 | 0.00 |
| cdkn2a | 13.73 | 1 | 0.00 |
| mycn | 3.90 | 1 | 0.05 |
| nf1 | 5.38 | 1 | 0.02 |
| met | 16.80 | 1 | 0.00 |
| TOTAL | 172.59 | 13 | 0.00 |

Table S1. Wald test for coefficients in the Gene Enhanced 6 Model

##### 2.1.2 Hazard and Hazard Ratios

| Variable | Low Value | High Value | Diff. | Effect | S.E. | Lower 0.95 | Upper 0.95 |
|---|---|---|---|---|---|---|---|
| age | 3.13 | 11.36 | 8.23 | 0.07 | 0.19 | -0.29 | 0.44 |
| Hazard Ratio | 3.13 | 11.36 | 8.23 | 1.07 | NA | 0.75 | 1.55 |
| risk_group - low:intermediate | 2.00 | 1.00 | NA | -0.28 | 0.19 | -0.65 | 0.09 |
| Hazard Ratio | 2.00 | 1.00 | NA | 0.75 | NA | 0.52 | 1.09 |
| risk_group - high:intermediate | 2.00 | 3.00 | NA | 0.88 | 0.16 | 0.56 | 1.20 |
| Hazard Ratio | 2.00 | 3.00 | NA | 2.41 | NA | 1.75 | 3.31 |
| fusion - fp:fn | 1.00 | 2.00 | NA | 0.88 | 0.18 | 0.54 | 1.23 |
| Hazard Ratio | 1.00 | 2.00 | NA | 2.42 | NA | 1.72 | 3.42 |
| cohort - uk:cog | 1.00 | 2.00 | NA | 0.37 | 0.14 | 0.10 | 0.64 |
| Hazard Ratio | 1.00 | 2.00 | NA | 1.45 | NA | 1.11 | 1.91 |
| myod1 - y:n | 1.00 | 2.00 | NA | 1.52 | 0.32 | 0.88 | 2.15 |
| Hazard Ratio | 1.00 | 2.00 | NA | 4.57 | NA | 2.42 | 8.62 |
| tp53 - y:n | 1.00 | 2.00 | NA | 1.00 | 0.19 | 0.63 | 1.37 |
| Hazard Ratio | 1.00 | 2.00 | NA | 2.72 | NA | 1.88 | 3.94 |
| cdkn2a - y:n | 1.00 | 2.00 | NA | 1.16 | 0.31 | 0.55 | 1.77 |
| Hazard Ratio | 1.00 | 2.00 | NA | 3.18 | NA | 1.72 | 5.87 |
| mycn - y:n | 1.00 | 2.00 | NA | 0.68 | 0.34 | 0.00 | 1.36 |
| Hazard Ratio | 1.00 | 2.00 | NA | 1.98 | NA | 1.00 | 3.88 |

| Variable | Low Value | High Value | Diff. | Effect | S.E. | Lower 0.95 | Upper 0.95 |
|---|---|---|---|---|---|---|---|
| nf1 - y:n | 1.00 | 2.00 | NA | -0.59 | 0.26 | -1.09 | -0.09 |
| Hazard Ratio | 1.00 | 2.00 | NA | 0.55 | NA | 0.34 | 0.91 |
| met - y:n | 1.00 | 2.00 | NA | 1.78 | 0.43 | 0.93 | 2.63 |
| Hazard Ratio | 1.00 | 2.00 | NA | 5.91 | NA | 2.53 | 13.83 |

Table S2. Hazard Ratios and log(Hazards) (rows above rows labeled 'Hazard Ratios') for the Gene Enhanced 6 Model

#### 2.2 Gene Enhanced 2 Model

##### 2.2.1 Coefficients

| Variable | Chi-Square | d.f. | P |
|---|---|---|---|
| age | 11.57 | 3 | 0.01 |
| Nonlinear | 5.79 | 2 | 0.06 |
| risk_group | 38.18 | 2 | 0.00 |
| fusion | 15.23 | 1 | 0.00 |
| cohort | 3.12 | 1 | 0.08 |
| TOTAL | 104.51 | 7 | 0.00 |

Table S3. Wald test for coefficients in the Gene Enhanced 2 Model

##### 2.2.2 Hazard and Hazard Ratios

| Variable | Low Value | High Value | Diff. | Effect | S.E. | Lower 0.95 | Upper 0.95 |
|---|---|---|---|---|---|---|---|
| age | 3.13 | 11.36 | 8.23 | 0.14 | 0.18 | -0.21 | 0.50 |
| Hazard Ratio | 3.13 | 11.36 | 8.23 | 1.15 | NA | 0.81 | 1.65 |
| risk_group - low:intermediate | 2.00 | 1.00 | NA | -0.45 | 0.18 | -0.81 | -0.09 |
| Hazard Ratio | 2.00 | 1.00 | NA | 0.64 | NA | 0.44 | 0.92 |
| risk_group - high:intermediate | 2.00 | 3.00 | NA | 0.75 | 0.16 | 0.44 | 1.06 |
| Hazard Ratio | 2.00 | 3.00 | NA | 2.12 | NA | 1.55 | 2.90 |
| fusion - fp:fn | 1.00 | 2.00 | NA | 0.61 | 0.16 | 0.31 | 0.92 |
| Hazard Ratio | 1.00 | 2.00 | NA | 1.85 | NA | 1.36 | 2.52 |

| Variable | Low Value | High Value | Diff. | Effect | S.E. | Lower 0.95 | Upper 0.95 |
|---|---|---|---|---|---|---|---|
| cohort - uk:cog | 1.00 | 2.00 | NA | 0.24 | 0.13 | -0.03 | 0.50 |
| Hazard Ratio | 1.00 | 2.00 | NA | 1.27 | NA | 0.97 | 1.64 |

Table S4. Hazard Ratios and log(Hazards) (rows above rows labeled 'Hazard Ratios') for the Gene Enhanced 2 Model

#### 2.3 Baseline Clinical Model

##### 2.3.1 Coefficients

| Variable | Chi-Square | d.f. | P |
|---|---|---|---|
| age | 11.57 | 3 | 0.01 |
| Nonlinear | 5.79 | 2 | 0.06 |
| risk_group | 38.18 | 2 | 0.00 |
| fusion | 15.23 | 1 | 0.00 |
| cohort | 3.12 | 1 | 0.08 |
| TOTAL | 104.51 | 7 | 0.00 |

Table S5. Wald test for coefficients in the Baseline Clinical Model

##### 2.3.2 Hazard and Hazard Ratios

| Variable | Low Value | High Value | Diff. | Effect | S.E. | Lower 0.95 | Upper 0.95 |
|---|---|---|---|---|---|---|---|
| age | 3.13 | 11.36 | 8.23 | 0.14 | 0.18 | -0.21 | 0.50 |
| Hazard Ratio | 3.13 | 11.36 | 8.23 | 1.15 | NA | 0.81 | 1.65 |
| risk_group - low:intermediate | 2.00 | 1.00 | NA | -0.45 | 0.18 | -0.81 | -0.09 |
| Hazard Ratio | 2.00 | 1.00 | NA | 0.64 | NA | 0.44 | 0.92 |
| risk_group - high:intermediate | 2.00 | 3.00 | NA | 0.75 | 0.16 | 0.44 | 1.06 |
| Hazard Ratio | 2.00 | 3.00 | NA | 2.12 | NA | 1.55 | 2.90 |
| fusion - fp:fn | 1.00 | 2.00 | NA | 0.61 | 0.16 | 0.31 | 0.92 |
| Hazard Ratio | 1.00 | 2.00 | NA | 1.85 | NA | 1.36 | 2.52 |
| cohort - uk:cog | 1.00 | 2.00 | NA | 0.24 | 0.13 | -0.03 | 0.50 |

| Variable | Low Value | High Value | Diff. | Effect | S.E. | Lower 0.95 | Upper 0.95 |
|---|---|---|---|---|---|---|---|
| Hazard Ratio | 1.00 | 2.00 | NA | 1.27 | NA | 0.97 | 1.64 |

Table S6. Hazard Ratios and log(Hazards) (rows above rows labeled 'Hazard Ratios') for the Baseline Clinical Model
